# Interventions used to improve air flow in hospitals – a rapid review

**DOI:** 10.1101/2023.10.06.23296654

**Authors:** Gráinne Brady, Fiona Bennin, Rosaline De Koning, Manish K Tiwari, Cecilia Vindrola-Padros, Danielle Morris, Elizabeth Lloyd-Dehler, Jerry Slann, Simon Watt, Fiona Stevenson, Zarnie Khadjesari, Hakim-Moulay Dehbi, Andrea Ducci, Ryo Torii, Lena Ciric, Ruth Epstein, John Rubin, Catherine Houlihan, Rachael Hunter, Laurence B Lovat

**Affiliations:** Rapid Research Evaluation and Appraisal Lab (RREAL), Department of Targeted Intervention, University College London, UK; Department of Mechanical Engineering, University College London, UK; WEISS Centre, University College London, UK; East and North Hertfordshire NHS Trust, UK; Lay member; Institute of Occupational Medicine, UK; Institute of Epidemiology and Health Care, University College London, UK; School of Health Sciences, University of East Anglia, UK; Comprehensive Clinical Trials Unit, University College London, UK; Healthy Infrastructure Research Group, Department of Civil, Environmental and Geomatic Engineering, University College London, UK; Royal National Throat Nose and Ear Hospital, University College London Hospitals NHS Foundation Trust, London, UK; Department of Virology, University College London Hospitals NHS Foundation Trust, London, UK; Institute of Epidemiology & Health, Faculty of Population Health Sciences, University College London, UK; Division of Surgery and Interventional Science, University College London, UK

**Author notes:** Rapid Research Evaluation and Appraisal Lab (RREAL) Department of Targeted Intervention University College London (UCL) Charles Bell House 43-45 Foley Street W1W 7TY @RREALwork https://rapidresearchandevaluation.com/. Contact: Prof Cecilia Vindrola.

## Abstract

The COVID-19 pandemic has highlighted the need for improved air flow in hospitals, to reduce the transmission of airborne infections such as COVID-19. The aim of this review was to map the existing literature on intervention used to improve air flow in hospitals, understanding challenges in implementation and the findings of any evaluations. We reviewed peer-reviewed articles identified on three databases, MEDLINE, Web of Science and the Cochrane Library with no restriction on date. 5846 articles were identified, 130 were reviewed and 18 were included: ten articles were from databases and eight articles were identified through hand searching. Results were discussed in terms of three categories: (i) concentration of aerosol particles, (ii) changes in/effect of air speed and ventilation and (iii) improvements or reduction in health conditions. Eight studies included an evaluation, the majority only had one comparator condition however three had multiple conditions. The most common device or method that was outlined by researchers was HEPA filters, which can remove particles with a size of 3 microns. Articles outline different interventions to improve air flow and some demonstrate their effectiveness in terms of improving health outcomes for patients, they also suggest either mechanical and natural ventilation are the best methods for dispersing particulate matter as well as perhaps two air cleaning units rather than one. With different methods comes different strengths and weaknesses however, the key finding is that air flow improvement measures reduce the likelihood of nosocomial infections.

## Introduction

Transmission of severe acute respiratory syndrome coronavirus 2 (SARS-CoV-2) can either be via respiratory droplets through coughs and sneezes usually over a range of 2 meters, formite exposure through touch of an infected surface to a person’s eyes, nose or mouth or through airborne transmission (CDC, 2020; Li et al, 2020; Derqui et al, 2023; Borak, 2020). In July 2020, the WHO reported airborne transmission of SARS-CoV-2 was a main transmission route, likely happening in healthcare settings and crowded indoor areas such as restaurants, fitness classes and choir practice (WHO, 2020b).

The COVID-19 pandemic highlighted that many hospital buildings were inadequate at providing effective infection prevention and control (BMA, 2023) and demonstrated that drastic changes in hospital ventilation and air flow would be required (Knight et al, 2022). Improvements in hospital air flow are not easily implemented, as these might require structural changes, the investment of resources and training staff on operation and safety (NHS England, 2023), particularly with bed side units. The aim of this review was to map the existing literature on interventions used to improve air flow in hospitals, understanding challenges in implementation and the findings of any evaluations.

### Design

The review was designed following the approach for rapid evidence reviews developed by Tricco et al (2017) with scope to incorporate relevant grey literature. The review followed a phased approach, which begins with a broad search strategy that is expanded with each round of searches. We followed the Preferred Reporting Items for Systematic Reviews and Meta-Analysis (PRISMA) statement to guide the review design and the reporting of the methods and findings.

The research questions guiding the review were:

RQ1: What are the types of interventions currently being used to improve air flow in hospitals?

RQ2: Have any of these been evaluated? If so, what are the main evaluation findings? RQ3: What are the main lessons learnt from the implementation of these interventions?

### Search strategy and approach

The search strategy was developed by researchers and relevant clinical colleagues. The first phase of the search strategy was broad and was run on general databases such as Google Scholar and PubMed. This led to the selection of a preliminary list of resources. This list was scanned for relevant key terms. The final search strategy is available in Appendix 1.

The search was not limited in any way other than to streamline outcomes, interventions, and the environment of the study. There was no date, language or location limitation, the inclusion criteria is available in Appendix 2. Final searches were conducted at the end of July 2023 on three databases (MEDLINE, Web of Science and The Cochrane Library).

### Document selection

The search results were imported into EndNote and duplicates were removed. Once this was complete all included references were imported into Rayyan for screening.

Following the initial screening of title and abstract, three researchers cross-checked 10% of exclusions against the inclusion criteria. The remaining publications that met the inclusion criteria were organised and allocated randomly between three researchers to facilitate full text screening.

### The following inclusion criteria were applied

Peer reviewed articles where interventions improving air flow are mentioned in the context of hospital settings, this could also be discussed in terms of a respiratory virus/infection.

No restriction on date, language, or study location.

To ensure the search was manageable, we did not include articles related to air flow in any other environment, PhD theses, dissertations, books, conference proceedings, incomplete versions, articles where we could not access the full text or letters to editors. The full inclusion criteria is available in Appendix 2.

### Data extraction

Data extraction was conducted using Microsoft Excel to organise the review process. The extraction form was piloted with two initial studies, and amendments were made before extracting data from all included studies. Data was extracted by three reviewers and checked by another member of the team.

### Data synthesis

Data were synthesised using narrative synthesis.

### Quality assessment

The methodological quality of the empirical articles was critically appraised using the Mixed Methods Appraisal Tool (MMAT) (Hong et al, 2018). The MMAT was developed to allow systematic reviewers assess the methodological quality of diverse study designs, including qualitative, quantitative, and mixed methods.

## Results

### Article selection

The initial search yielded 5848 articles (after duplicates were removed), 5715 articles were excluded as these did not meet the inclusion criteria outlined above, one article was removed at this stage as it could not be retrieved. We reviewed 130 articles at the full text stage and excluded 112 because they did not describe air flow, were not carried out in a hospital setting, were a simulation or model of air flow, or were excluded study designs such as reviews, or was a wrong publication type. 18 articles were included in the review (see Figure 1 for the PRISMA Flow Diagram).

**Figure 1.**
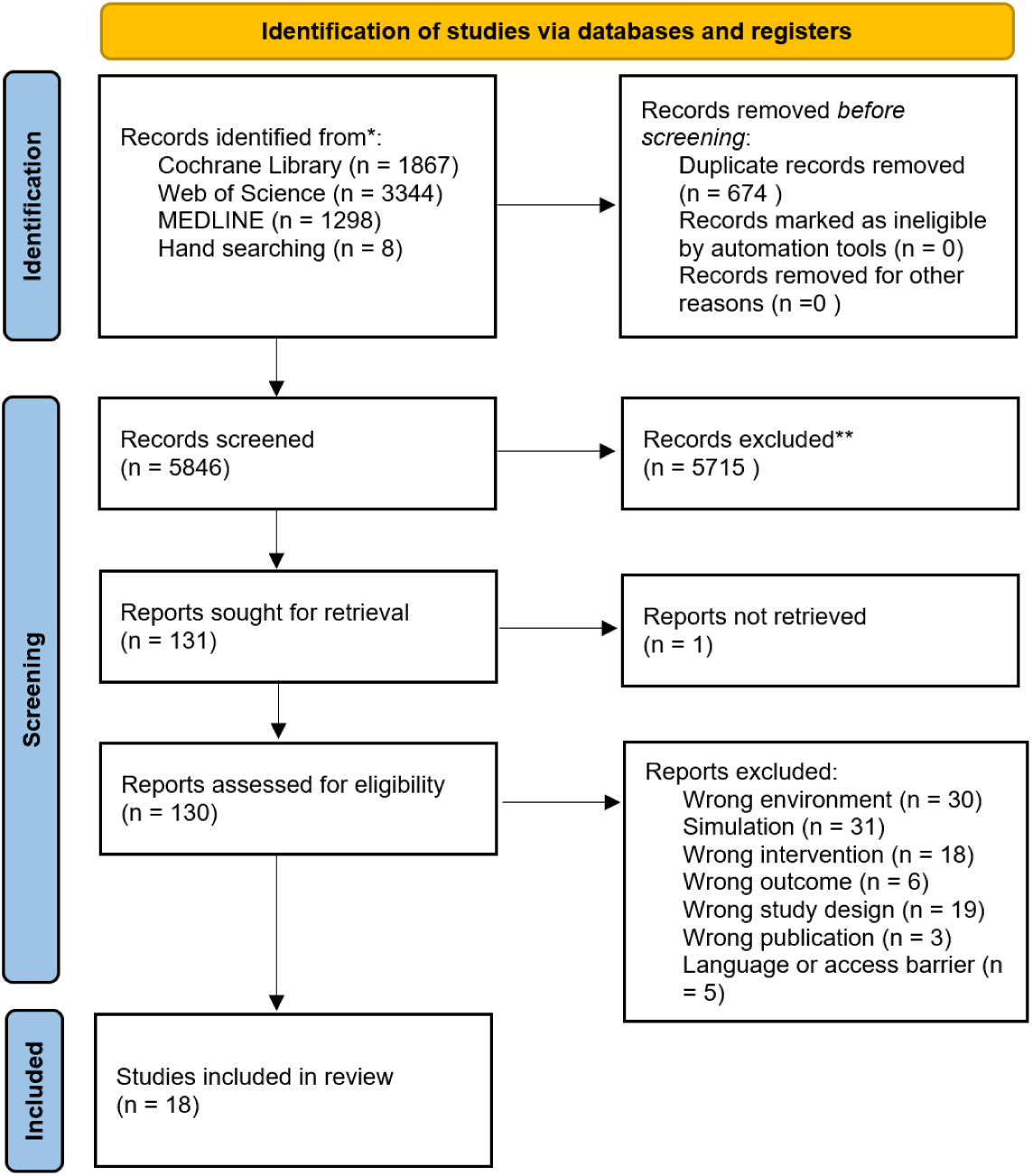
PRISMA Flow diagram.

### Article characteristics

Main article characteristics are summarised in Table 1. Three articles were from the UK, three from the USA, two from Australia, one article from China, Hong Kong, Ireland, Korea, Germany, Czech Republic, Singapore, South Africa, Iran and Italy.

**Table 1.**
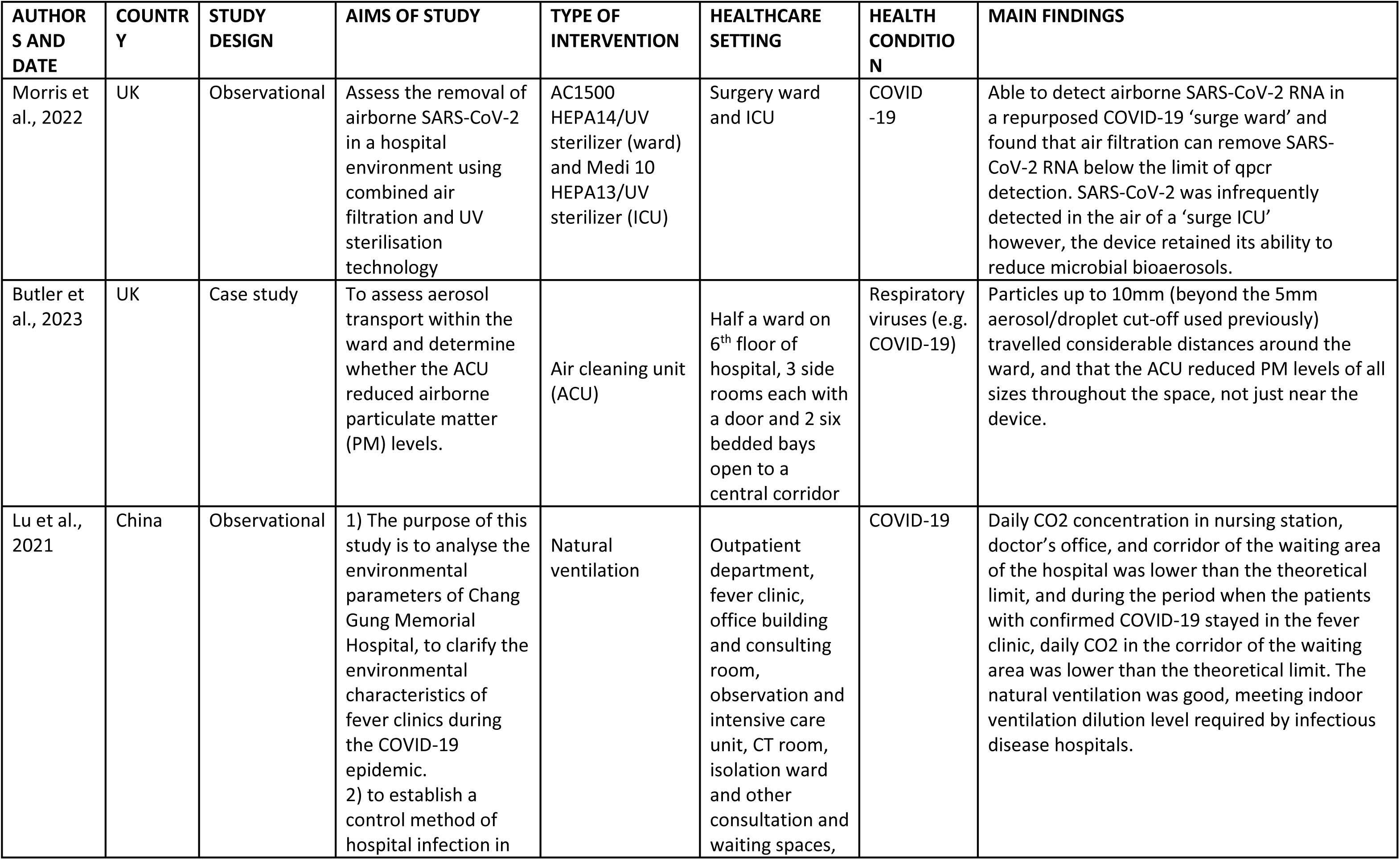
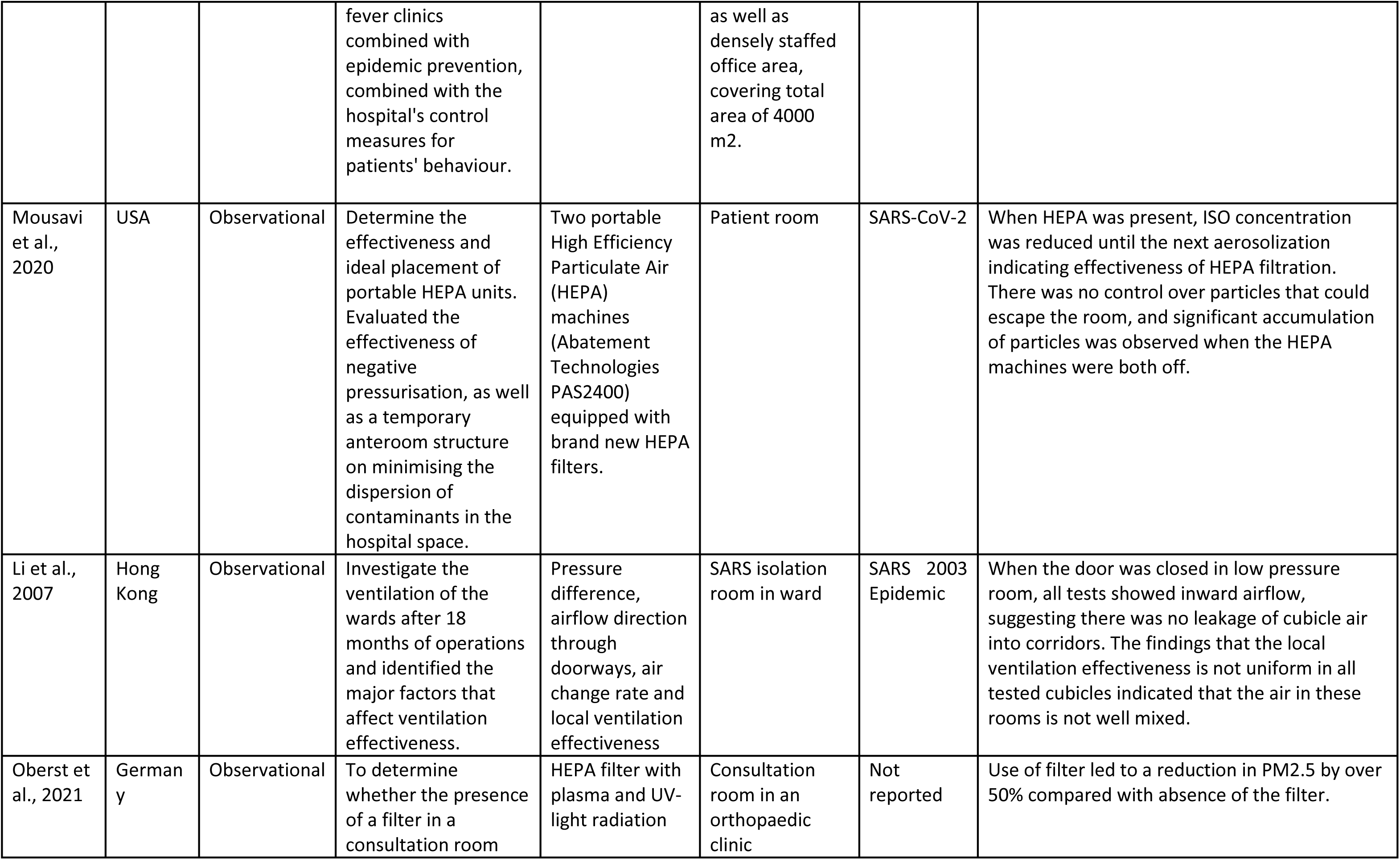
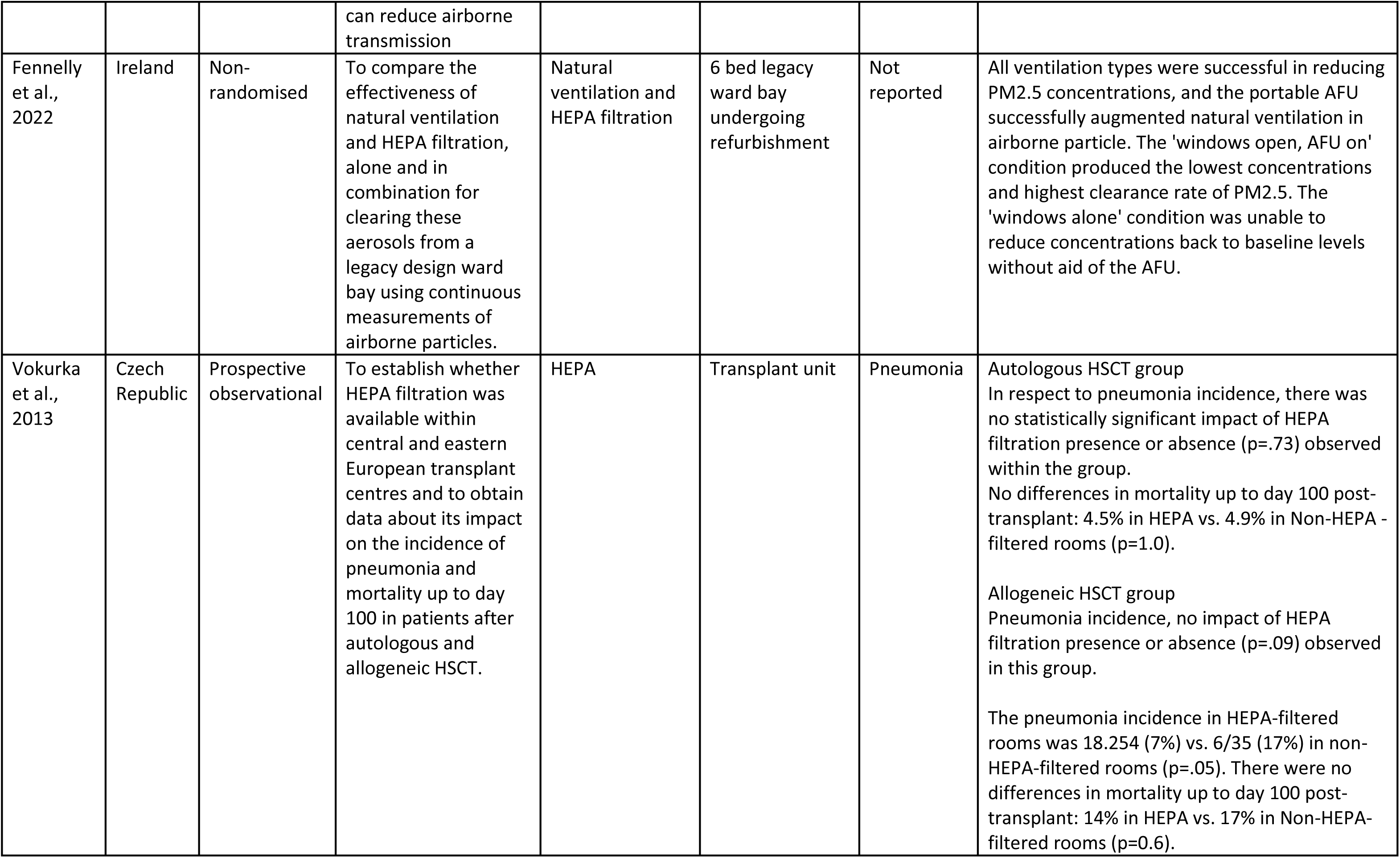
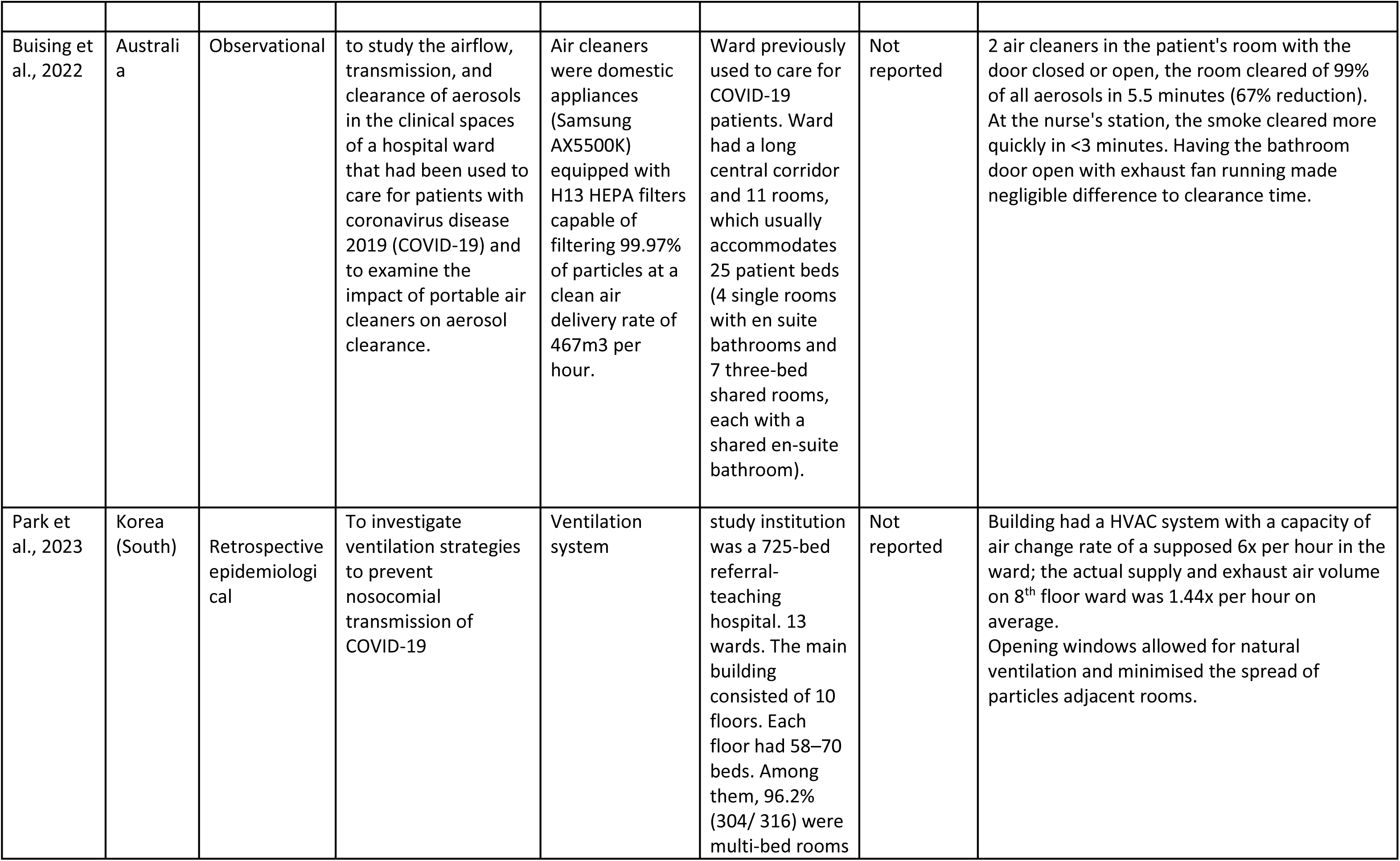
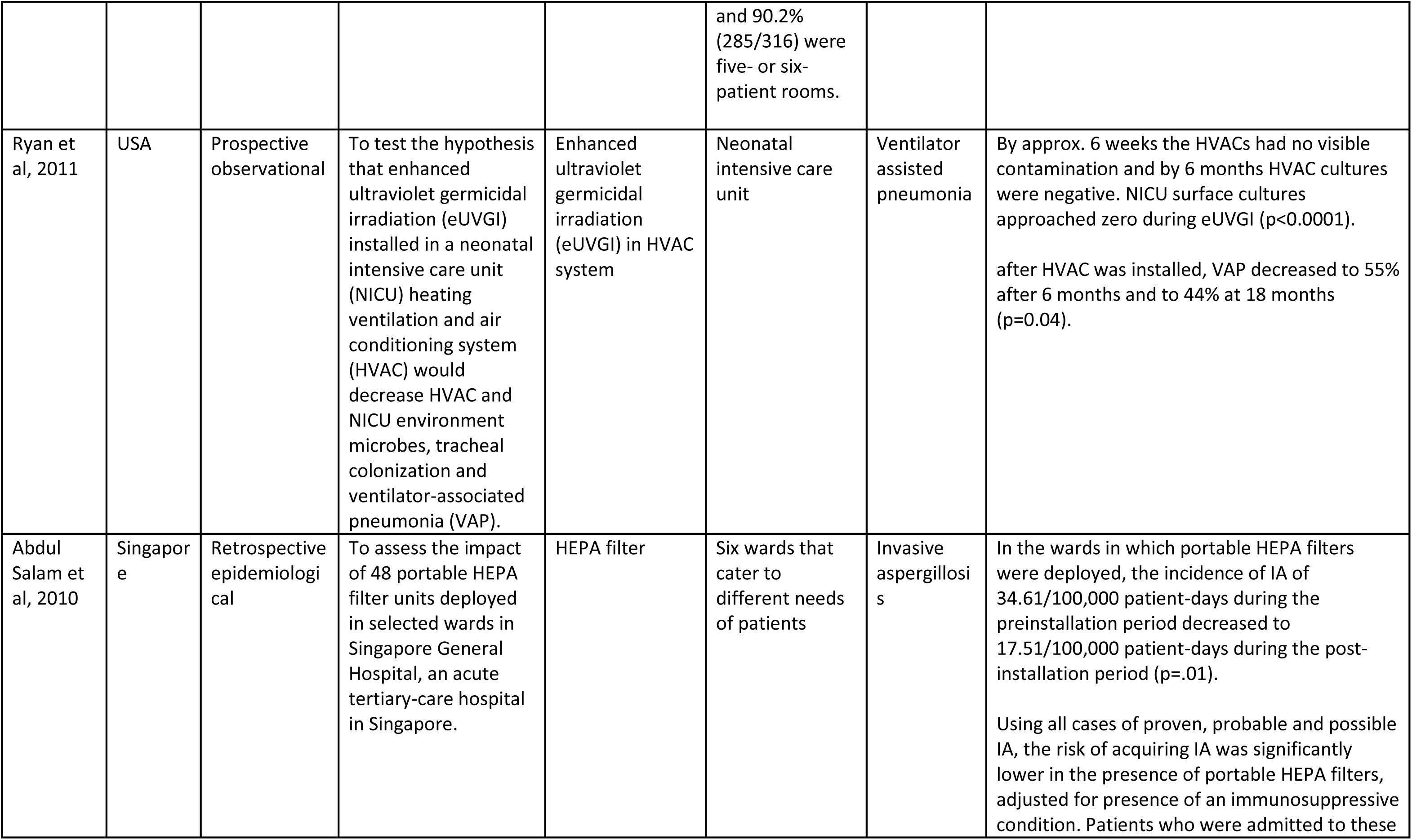
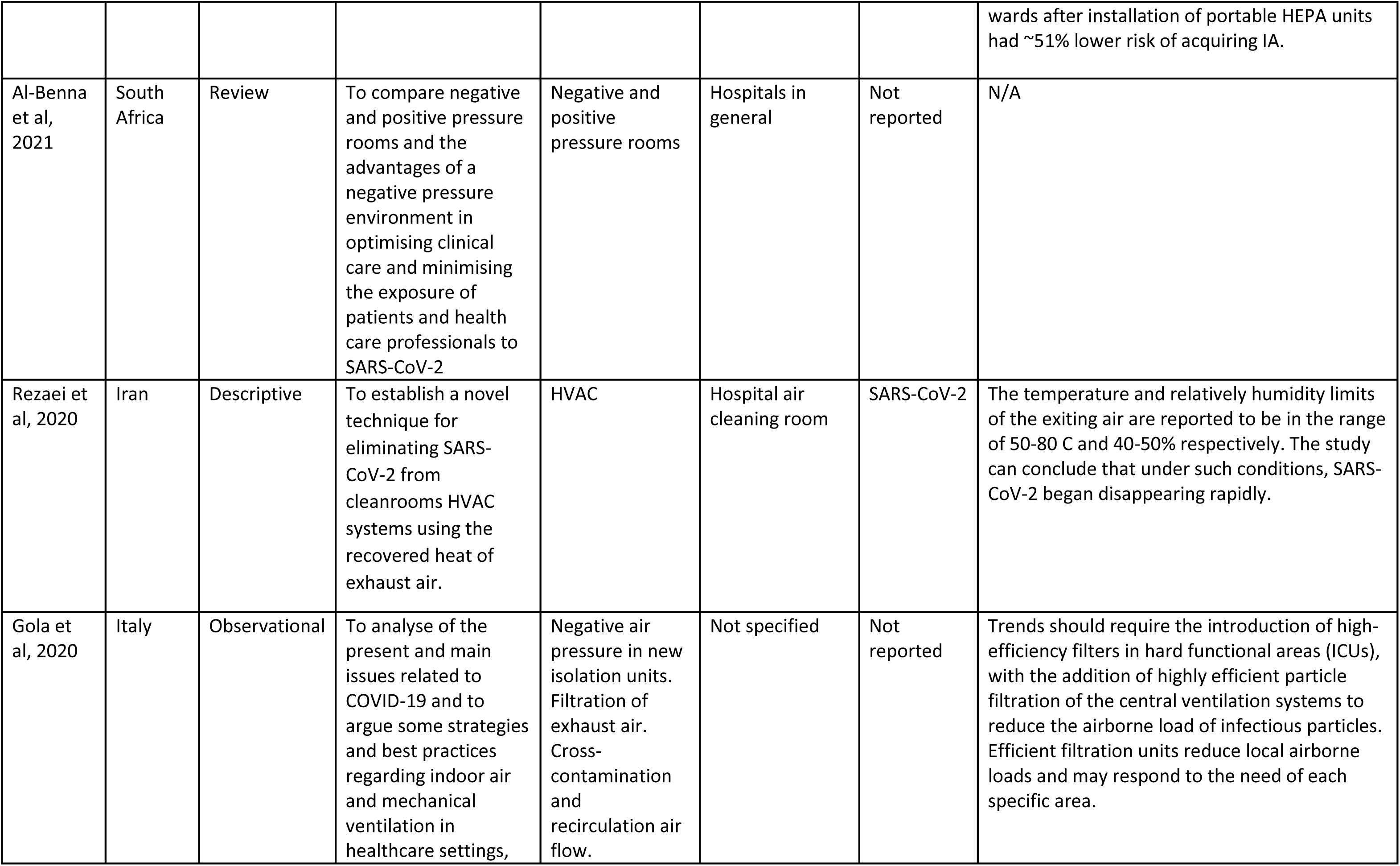
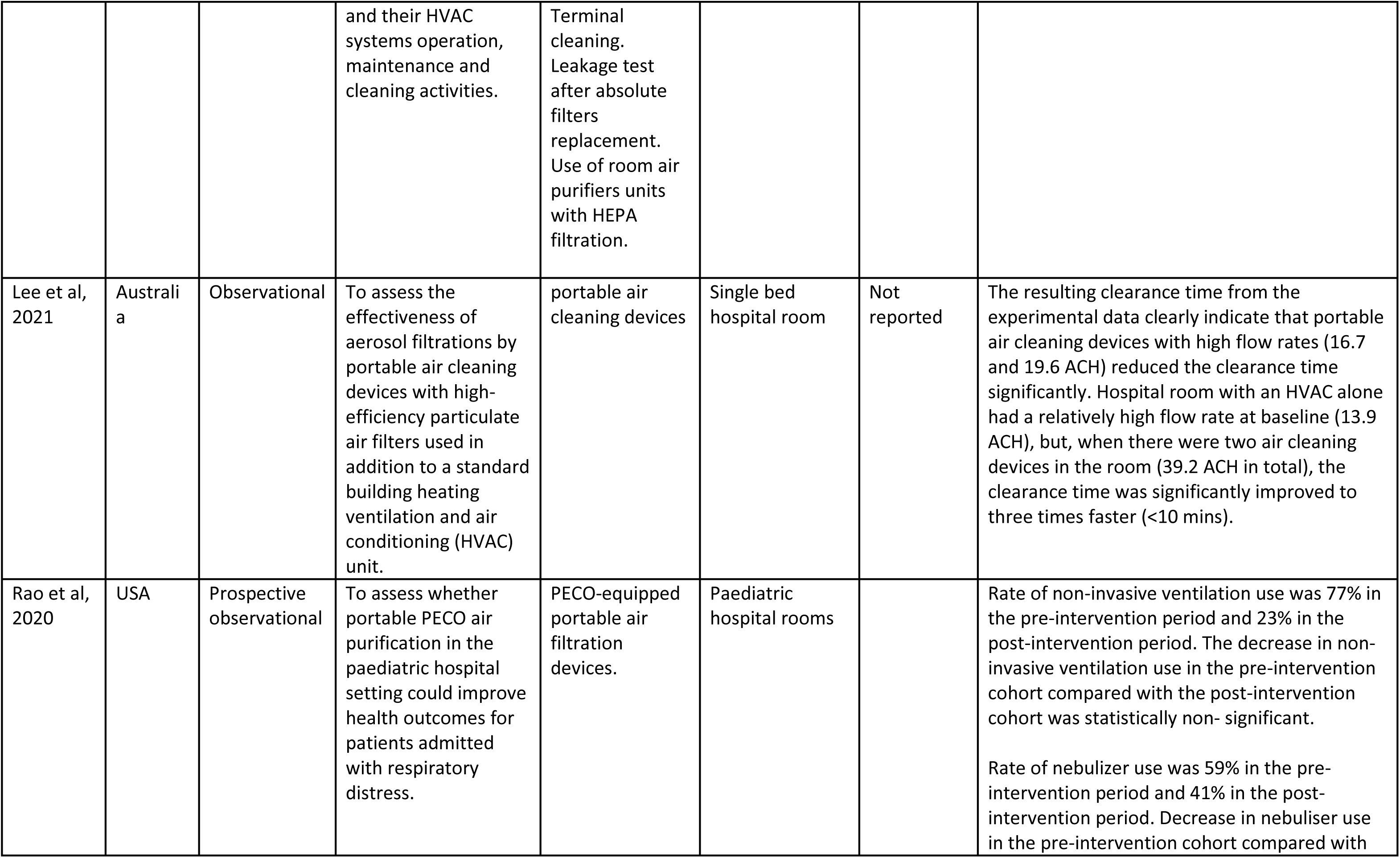
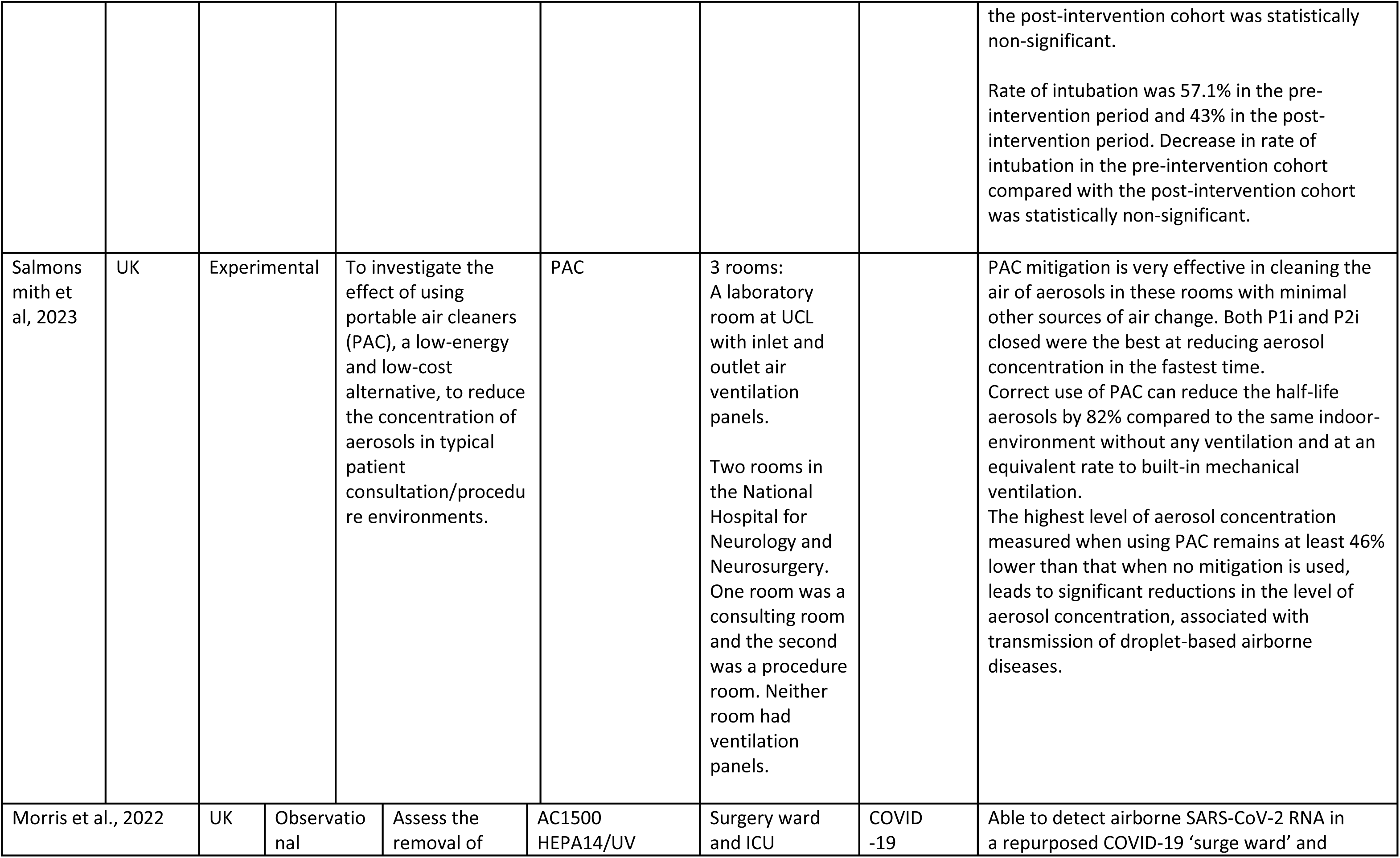
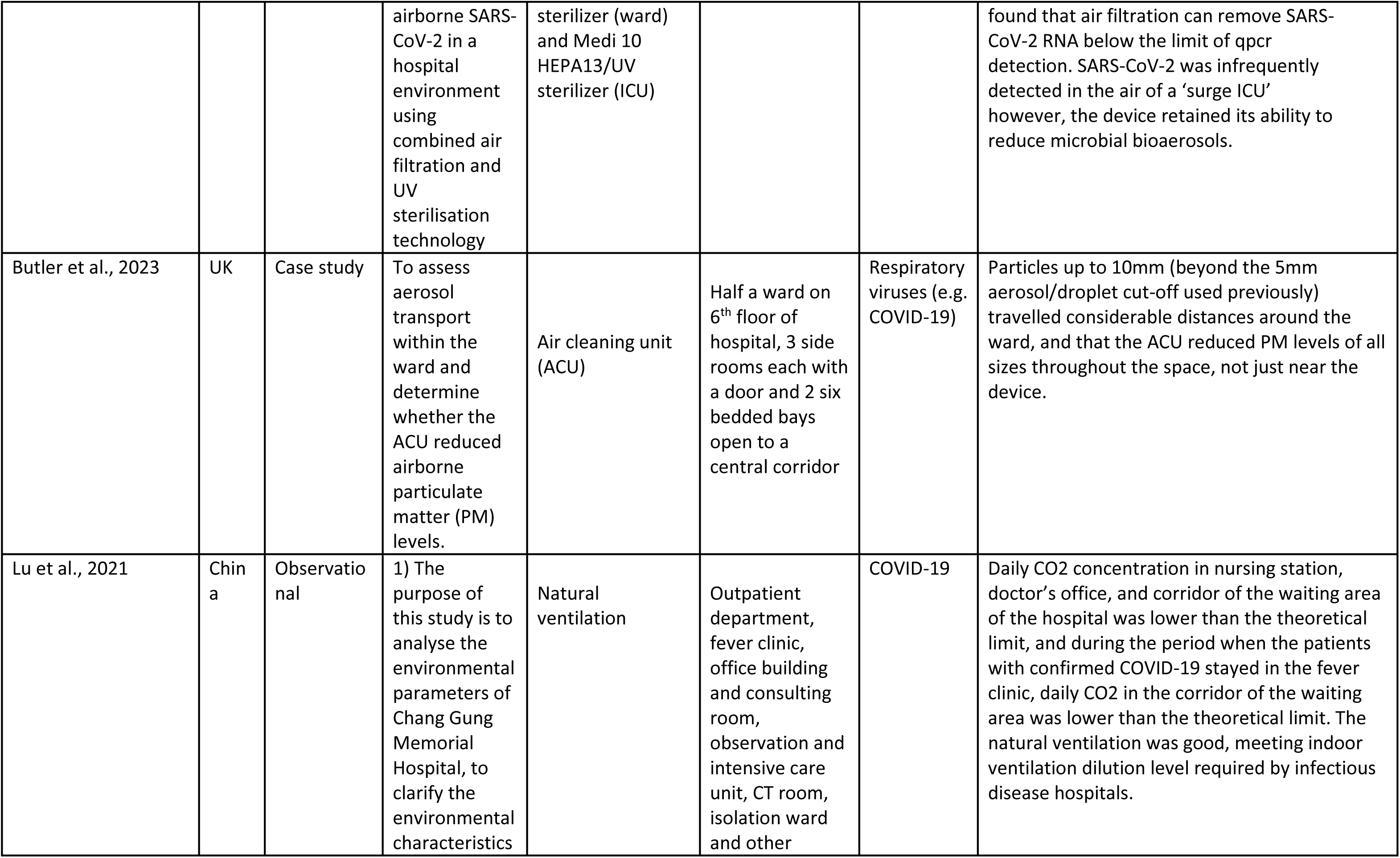
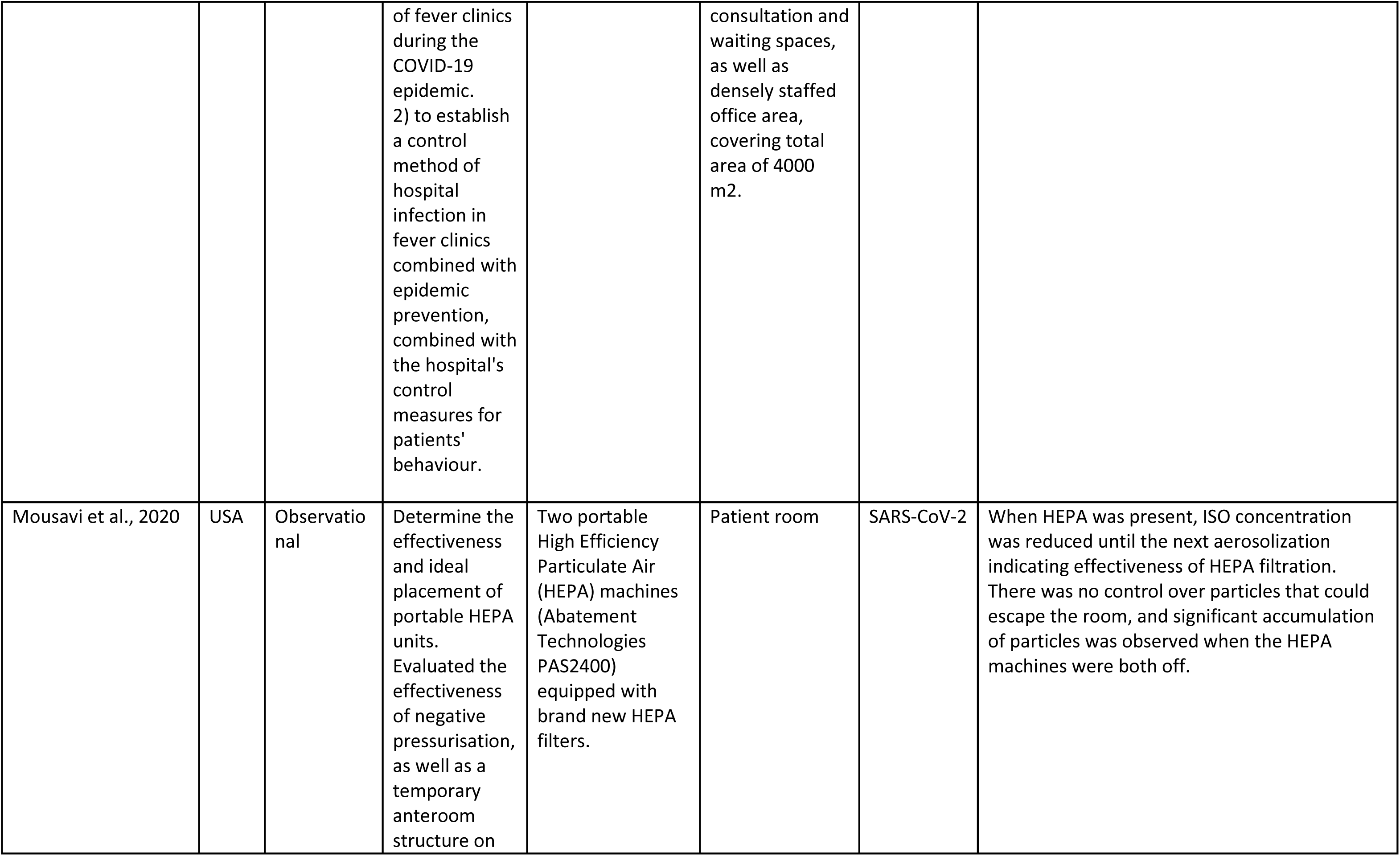
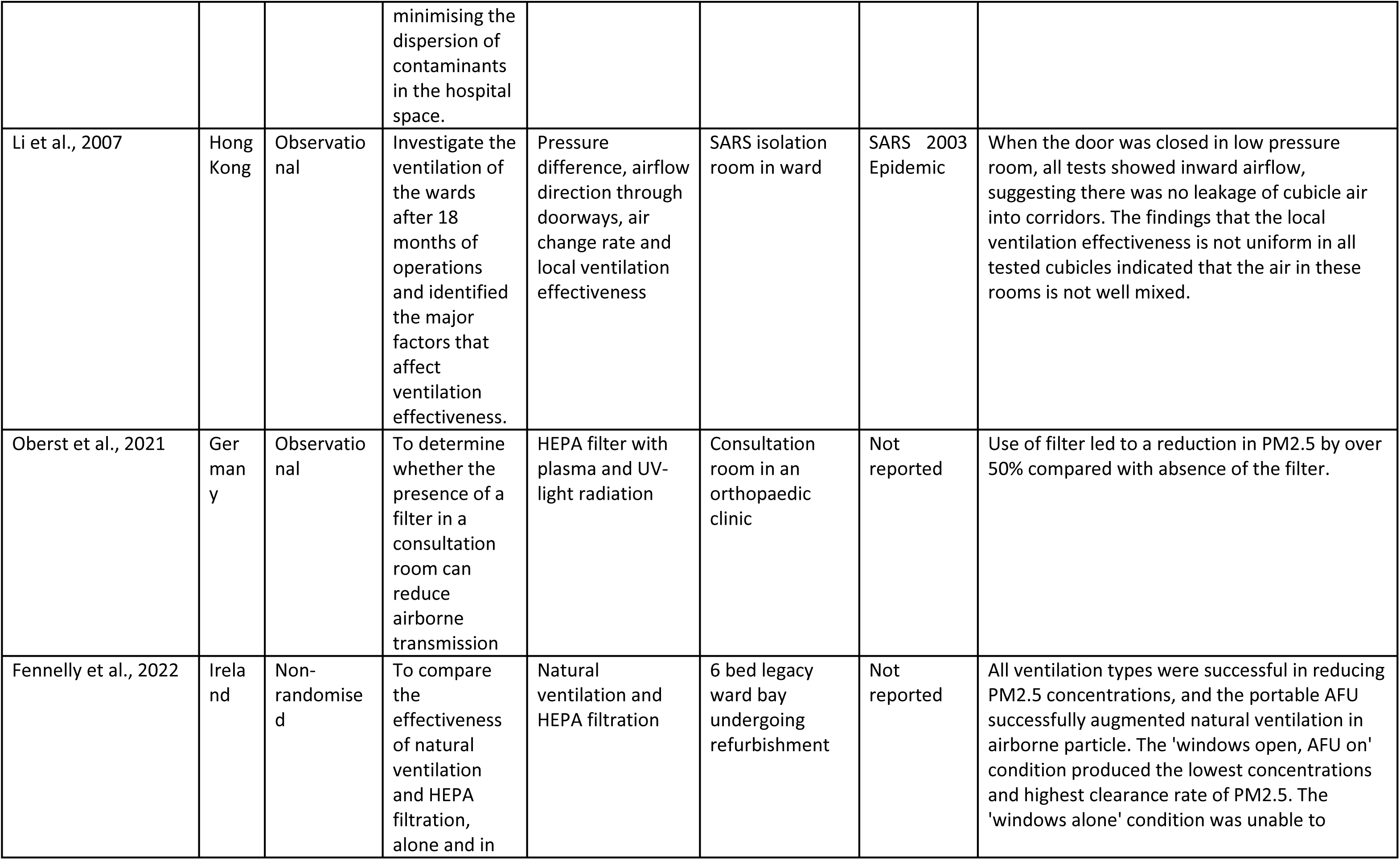
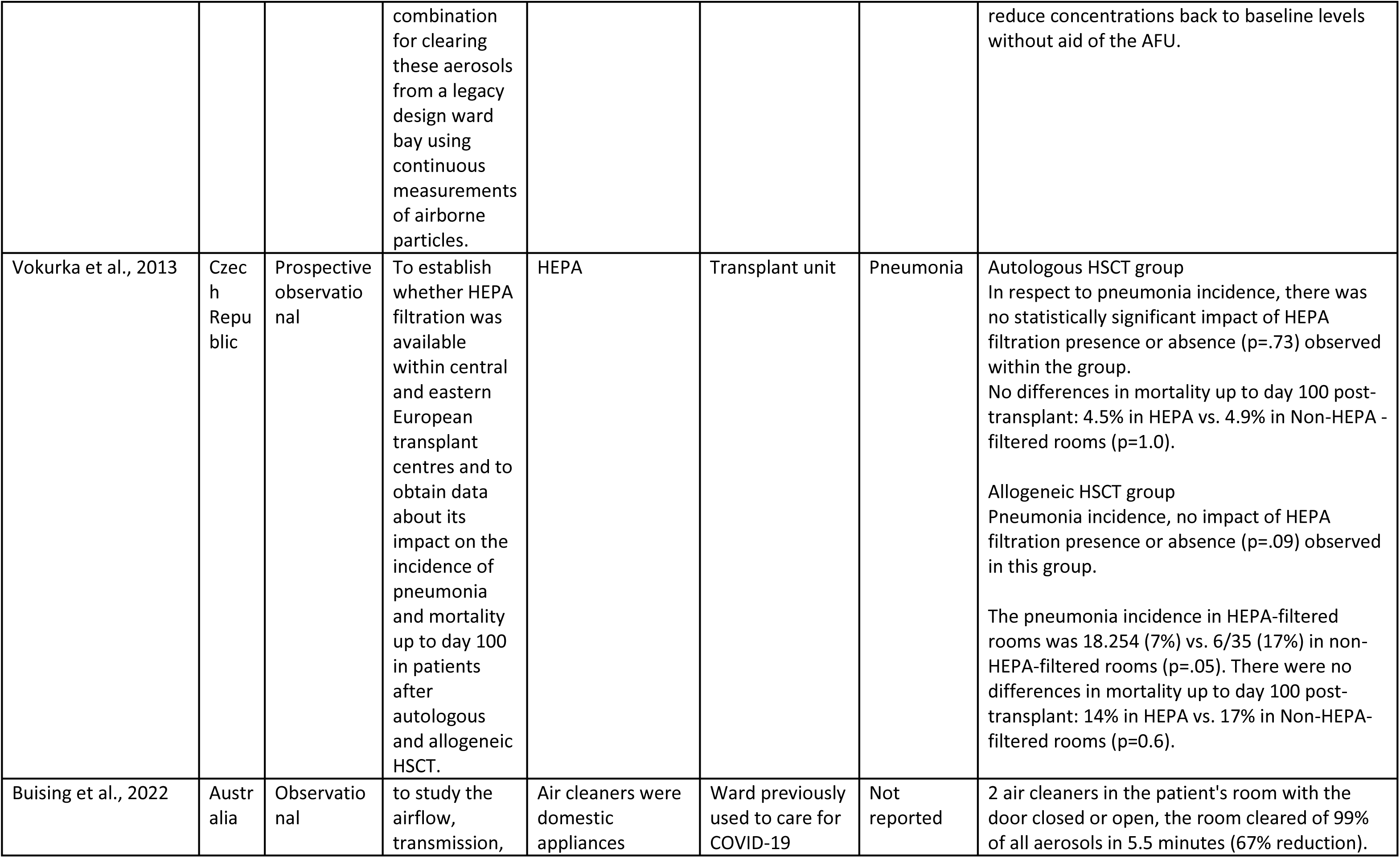
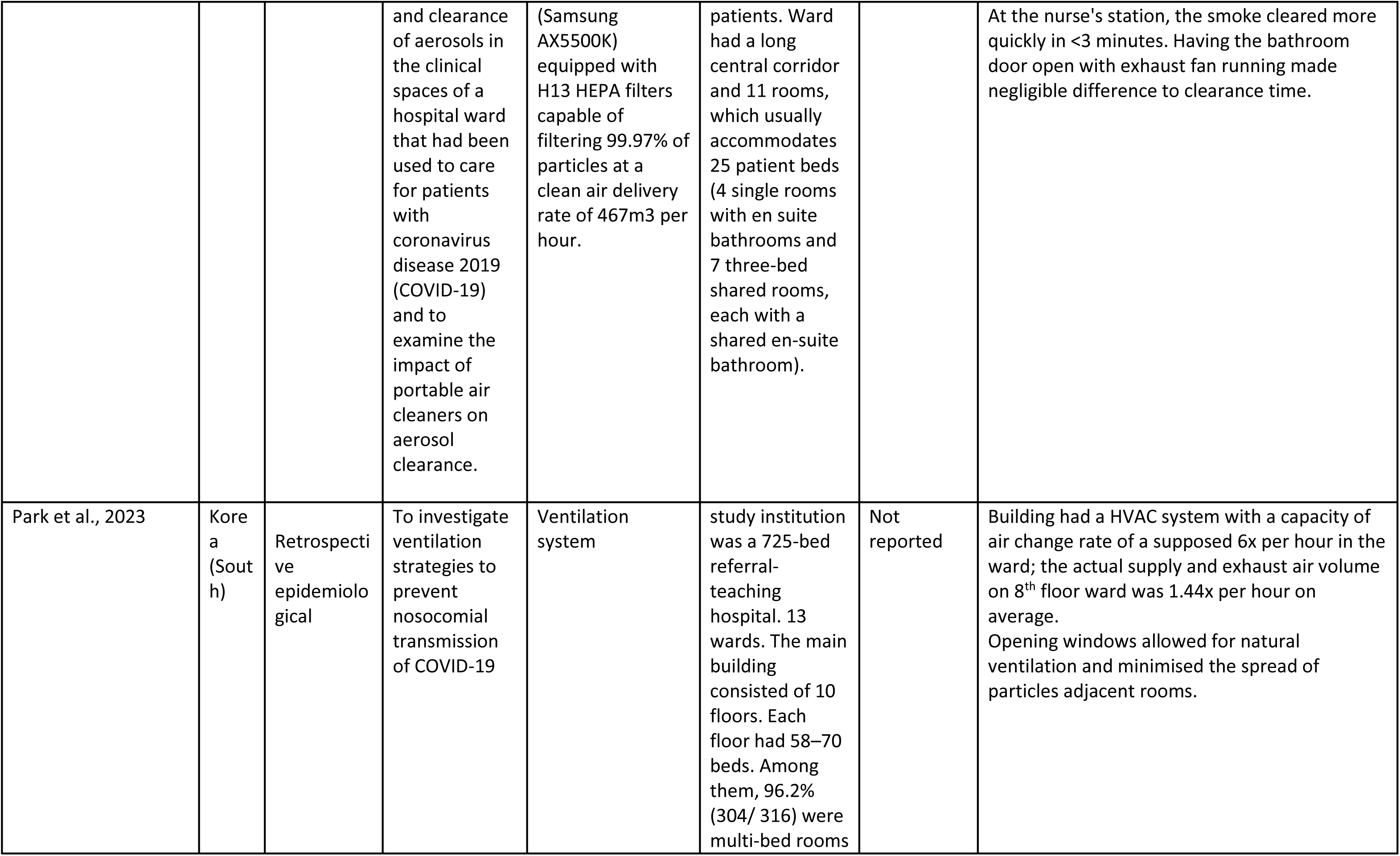
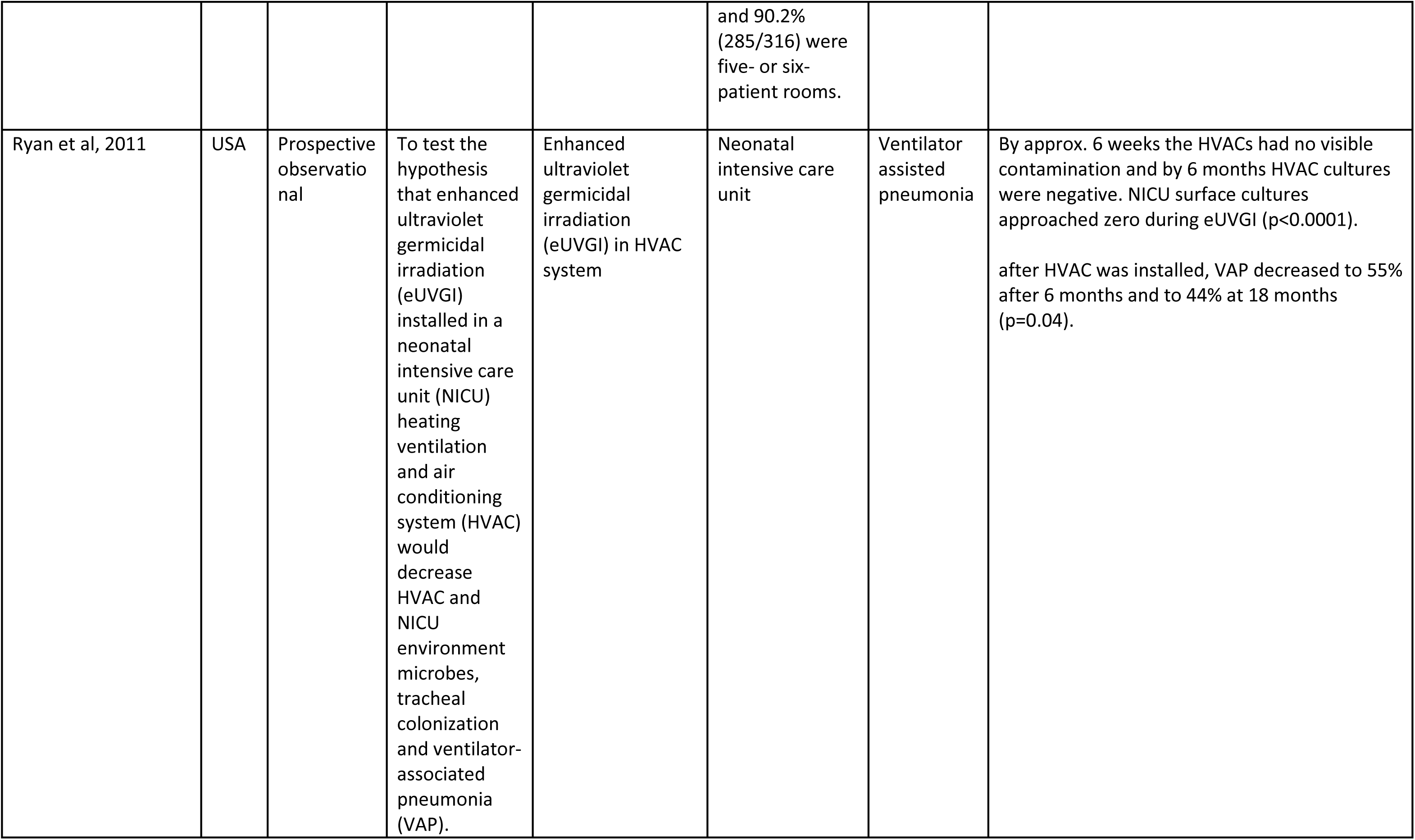
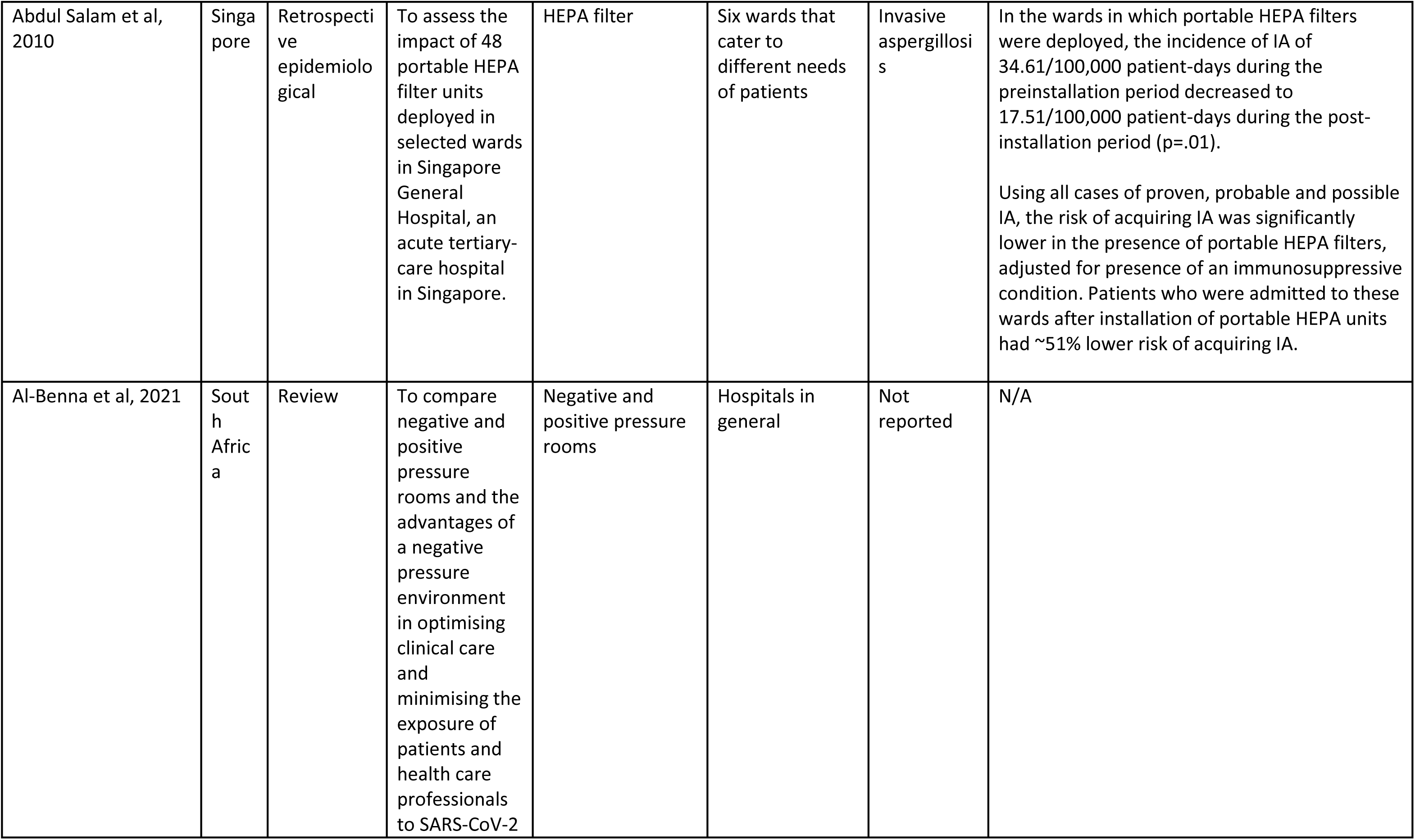
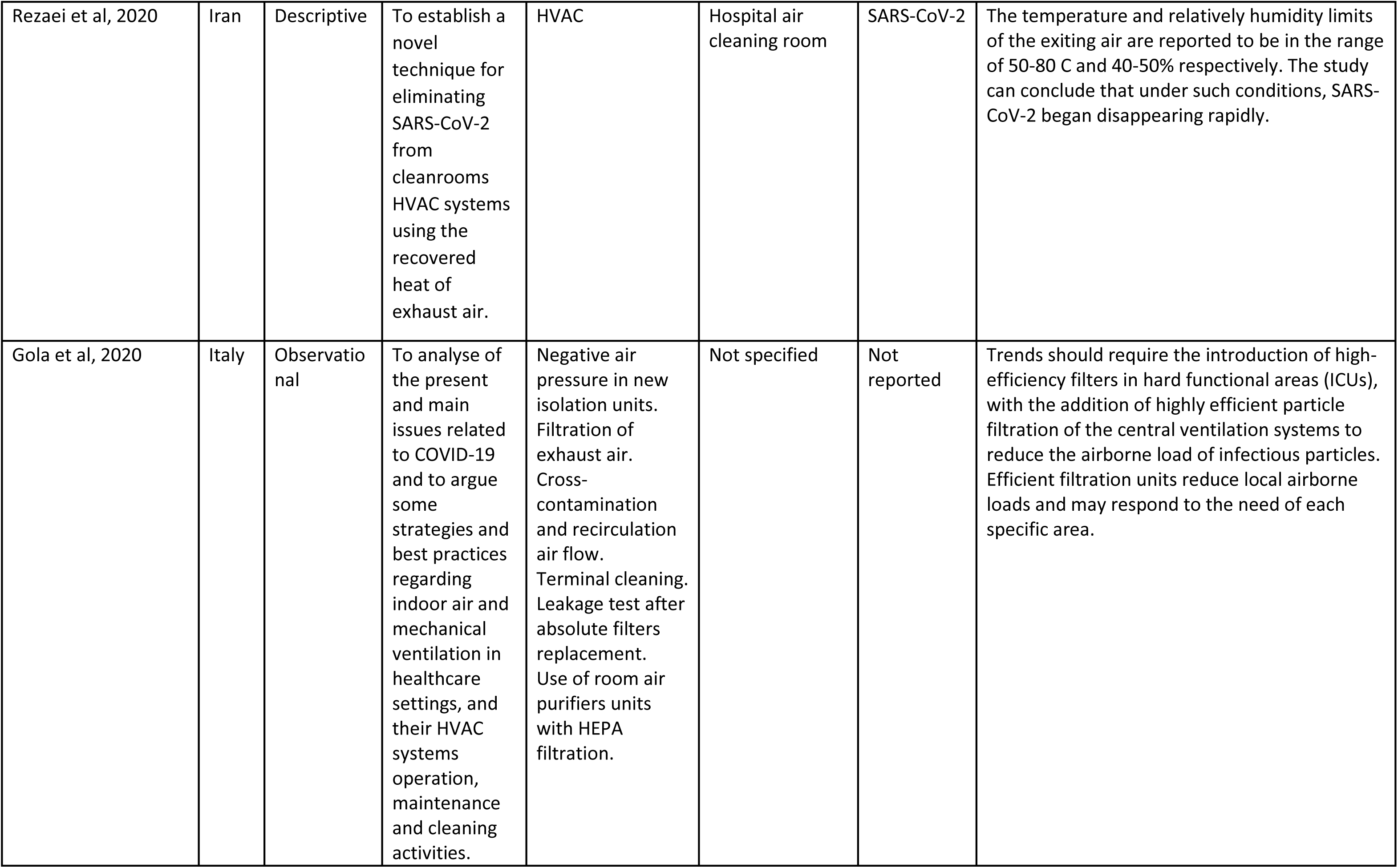
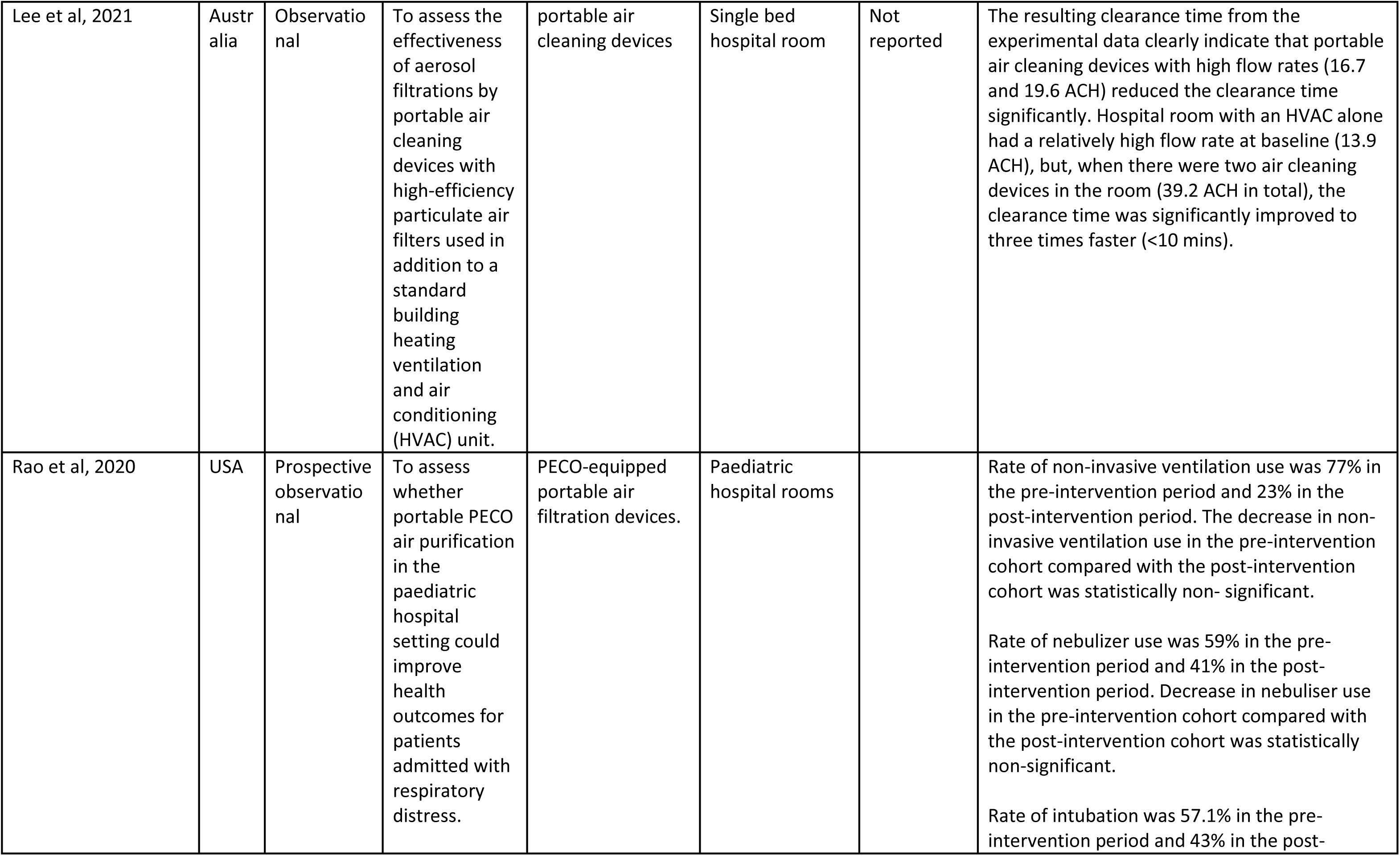
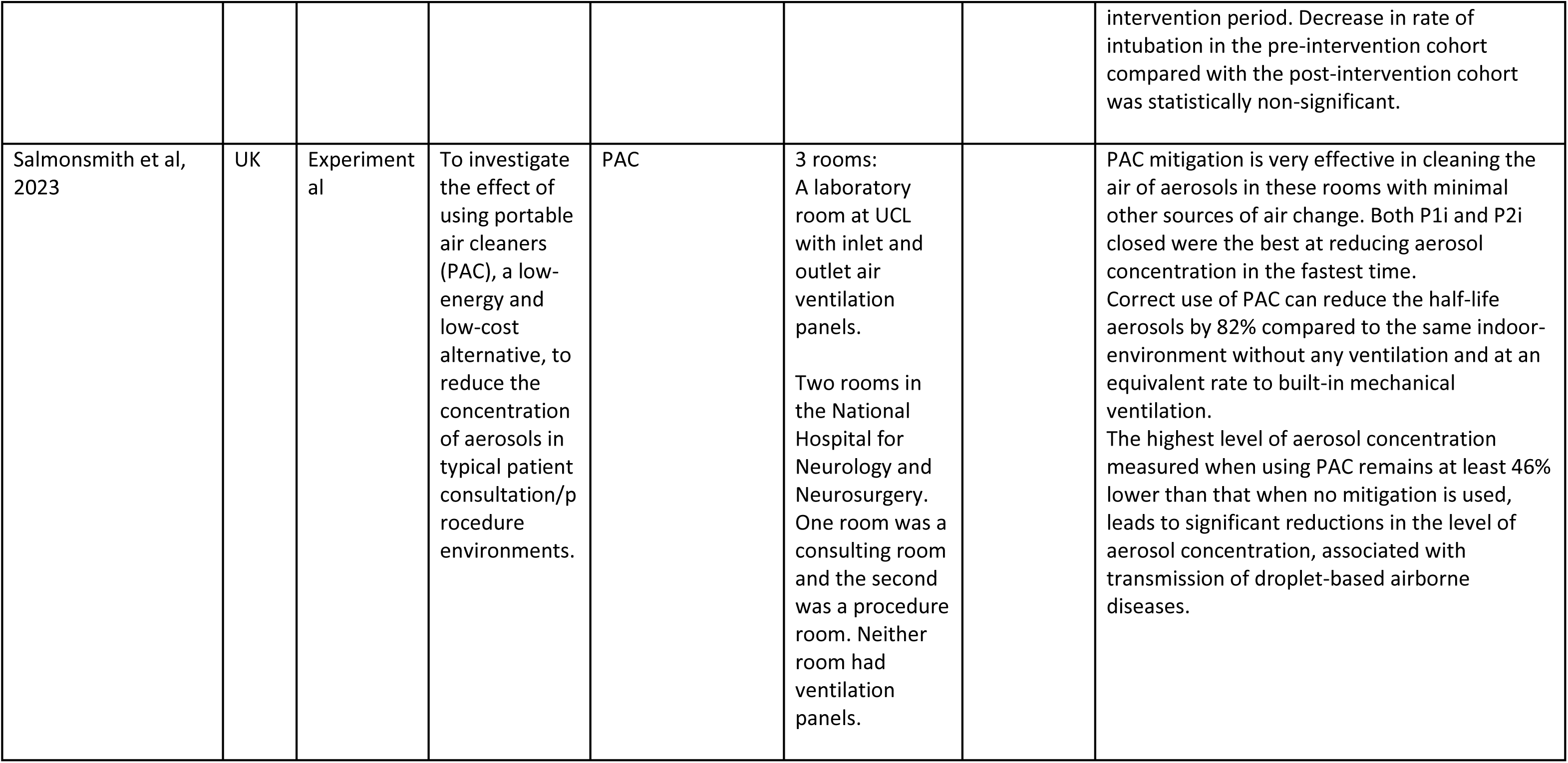
Study characteristics.

## Findings

The findings from the studies can be divided into three themes. The first theme includes findings around the concentration of aerosol particles and the second theme discusses changes in air speed and ventilation and the third theme is improvements or reductions of health conditions as a result of interventions.

## RQ1 & RQ3

All 18 articles outlined interventions that are currently used to improve air flow in hospitals.

### Concentration of aerosol particles

Conway Morris et al (2022) discussed the accumulation of aerosols depending on whether patients had respiratory support, suggesting that patients in ICU were commonly at a later stage of disease and as a result, may secrete fewer virus particles when they exhale. These findings suggest that aerosol precautions may, therefore, be more important in conventional wards than in well-defined aerosol risk areas.

Seven of the studies discussed the importance of ACUs, HEPA machines and filters in reducing the amount and size of aerosol particles in the air. Butler et al (2023) found that particles up to 10µm travelled considerable distances around a ward (beyond 2m), however, the ACU reduced the PM levels throughout the space (not just near the device). Oberst and Heinrich (2021) similarly reported that the addition of a filter into a consultation room could significantly reduce the risk of airborne transmission, with aerosol concentration decreasing by a minimum of 50%. Mousavi et al (2022) found that there was a significant accumulation of particles observed when the HEPA machines were both off, suggesting the key role of filtration in maintaining the air cleanliness. One study suggested that air ventilation (open windows) alone was unable to reduce concentrations back to baseline levels without the aid of an AFU, with the ‘windows open, AFU on’ condition producing the lowest concentrations and highest clearance rate of PM2.5 (Fennelly et al, 2022). Salmonsmith et al (2023) reported that PAC mitigation is very effective in clearing the air of aerosols in rooms, this could also reduce the half-life aerosols by 82%, Gola et al (2020) also found that filtration units reduced local airborne loads and suggested they should be used in hard functional areas such as ICUs. As Rezaei et al (2020) looked at cleaning rooms, they identified that a HVAC system provided exhaust air ranging from 50-80 degrees Celsius and with 40-50% humidity, under these conditions COVID-19 was observed to rapidly disappear improving infection control across whole hospitals.

Conversely, one study found that the existing ward HVAC system alone was inefficient when clearing a patient room of aerosols and that commercially available air cleaners may have a key role in clearing aerosolized particles that may contain respiratory viruses, such as SARS-CoV-2, in clinical environments (Buising et al, 2022).

### Changes in/effect of air speed and ventilation

When looking at measuring ventilation changes as a result of changes in the number of people in the room, it was found that, when there had been a failure to ‘dilute’ the air through ventilation, it was necessary to increase the ventilation rate (either naturally or through mechanical ventilation) or reduce the amount of people allowed in the room (Lu et al, 2021). When testing the movement of air in a sealed ward one study found that, when the door was closed, all tested wards had inward airflow or there was no outward airflow, suggesting that these new SARS wards are effective in securing no-leakage of cubicle air into the corridors, even when some of the cubicles failed to maintain a negative pressure difference (Li et al, 2007). Portable air cleaning units with high flow rates (16.7 & 19.6 ACH) reduced clearance time significantly, two ACUs in a room reduced the clearance time to under 10 minutes (Lee et al, 2021). Ryan et al (2011) looked at changes after introducing enhanced UV germicidal irradiation in HVAC systems, after six-weeks contamination on the system was not detected and at 6-months HVAC cultures were near zero (p<0.0001), showing the eUVGI to be extremely effective in reducing contamination.

One study found that, even though HEPA filtration and its regular monitoring were declared to be used, the authors could not collect objective data verifying its real efficacy and function on site (Vokurka et al, 2014). Similarly, Park et al (2023) found that even with a ventilation system, opening windows allowed for natural ventilation and minimised the spread of particles to adjacent rooms.

### Improvements or reduction in health conditions

Three articles outlined improvements in health conditions as a result of an intervention to improve air flow. Ryan et al (2011) reported after using an enhanced UV germicidal irradiation in HVAC systems, ventilator-assisted pneumonia (VAP) was decreased to 55% after 6-months and 44% after 18-months. Abdul-Salam et al (2010) found that after HEPA filters were installed, incidences of invasive aspergillosis (IA) significantly decreased, individuals admitted after the HEPA filters were installed had around 51% lower risk of acquiring IA. Rao et al (2020) found improvements after the implementation of portable PECO air purifiers. Pre to post-intervention for patients showed non-invasive ventilation improved from 77% to 23%, rate of nebuliser use from 59% to 41% and rate of intubation from 57.1% to 43%, however, these findings were not statistically significant.

## RQ2: Evaluations

A description of the intervention being evaluated (and whether a control condition has been used) and the evaluation findings are presented in table 2.

**Table 2.** Evaluations of interventions and main evaluation findings.

| Author | Description of Evaluation | Evaluation findings |
| --- | --- | --- |
| Morris et al, 2022 | Crossover evaluation, with the primary outcome being detection of SARS-CoV-2 RNA in the various size fractions of air samples. Differences in numbers of pathogens detected with filters on and off were compared. | Able to detect airborne SARS-CoV-2 RNA in a repurposed COVID-19 “surge ward” and found that air filtration can remove SARS-CoV-2 RNA below the limit of qPCR detection. SARS-CoV-2 was infrequently detected in the air of a “surge ICU”; however, the device retained its ability to reduce microbial bioaerosols. |
| Oberst & Heinrich, 2021 | The presence of aerosols was measured in the room both with the filter present and absent from the room during routine consultations. | Reduction in PM2.5 by over 50% when the filter was on. |
| Fennelly et al, 2022 | “Window alone condition. " three top hinged windows opened during this condition. | Highest calculated ACH was observed during the 'windows open, AFU on' condition. Mean PM2.5 clearance rate was significantly ($p<.01$) higher in the 'windows open, AFU on' condition compared with the 'AFU alone' condition, which, in turn, was significantly higher than the 'windows alone' conditions. Operation of the AFU, with or without open windows, reduced inter-monitor PM2.5 variations significantly. |
| Buising et al, 2022 | 1) Effect of installation of portable air cleaners in the patient room vs. usual HVAC on aerosols within and external to the room.<br>2) effect of patient room door open vs door closed on aerosols outside the room<br>3) effect of installation of variable number of portable air cleaners in the corridor on aerosols in the corridor<br>4) effect of installation of different barriers to enclose the nurse's station on aerosols in within the nurses' station | Using measurements within the patient room, at usual HVAC settings and with a closed door, it took 16 minutes for the aerosols in the patient room to clear back down to 1% of the baseline maximum measurable by the instruments. The relative amount of aerosols in the nurses' station compared to the patient room was highly variable representing 25%–50% of the room's concentration with the door closed using 2 air cleaners. When 2 air cleaners were placed in the patient room with the door closed or open, the room cleared of 99% of all aerosols in 5.5 minutes (a 67% reduction compared with no air cleaners). Having the bathroom door open with exhaust fan running made a negligible difference to the clearance time. |
| Li et al, 2007 | All isolation rooms could be operated in two modes, the SARS mode (i.e., operated with 100% fresh air) and the non-SARS mode (i.e., 70% air re-circulated). The study measured:<br>1. pressure difference, airflow<br>2. direction through doorways,<br>3. air change rate and local<br>4. ventilation effectiveness | 1 & 2): When the door was closed, all tested wards had inward airflow or there was no outward airflow, suggesting that these new SARS wards are effective in securing no-leakage of cubicle air into the corridors, even when some of the cubicles failed to maintain a negative pressure difference higher than 2.5 Pa.<br>3) Despite the fact that these SARS wards were designed nearly at the same time, with the state-of-the-art technologies, we still found that of the 35 tested cubicles, 26% had the air change rate less than 12 ACH, and all failed cubicles were from 3 of the 9 hospitals.<br>4) The findings that the local ventilation effectiveness was not uniform in all tested cubicles indicated that the air in these rooms was not well mixed. |
| Abdul-Salam et al, 2010 | Room(s) with HEPA filters vs. Room(s) without HEPA filters | No changes were seen in wards with no HEPA filtration, compared with improvements in cases of IA. |
| Lee et al, 2021 | Clean room<br>1) control room HVAC<br>2) Air cleaning device A<br>3) Air cleaning device B<br>4) Air cleaning device C<br>Hospital room<br>1) Hospital HVAC<br>2) 2x air cleaning device C and hospital HVAC | In the small control room, the aerosols were almost completely cleared 4-5x faster (<12 mins) with portable air cleaning devices than the control room with the HVAC system alone (2.3 ACH). The low-flow-rate portable air cleaning device with 8.3 ACH was still significantly better than the HVAC system alone. Hospital room with an HVAC alone had a relatively high flow rate at baseline (13.9 ACH), but, when there were two air cleaning devices in the room (39.2 ACH in total), the clearance time was significantly improved to three times faster (<10 mins). |
| Rao et al, 2020 | Pre-intervention vs. post-intervention | Rate of non-invasive ventilation use was 77% in the pre-intervention period and 23% in the post-intervention period. The decrease in non-invasive ventilation use in the pre-intervention cohort compared with the post-intervention cohort was statistically non-significant.<br>Rate of nebulizer use was 59% in the pre-intervention period and 41% in the post-intervention period. Decrease in nebuliser use in the pre-intervention cohort compared with the post-intervention cohort was statistically non-significant.<br>Rate of intubation was 57.1% in the pre-intervention period and 43% in the post-intervention period. Decrease in rate of intubation in the pre-intervention cohort compared with the post-intervention cohort was statistically non-significant. |

## MMAT findings

Two articles were not included in the quality appraisal as they were reviews and included to answer only RQ1. All five questions were not applicable for each study, therefore quality was assessed on the questions which are applicable, details are given in appendix 3. Seven articles were categorised as high quality, ranging from 80-100% of criteria met, five articles met criteria for medium quality (40-60%) and only one article met criteria for low quality at 25%. As this appraisal tool is mainly used to assess studies with human participants, the questions not applicable for our included studies were non-response bias and questions regarding sampling.

## Discussion

The aim of this rapid evidence review was to identify interventions being used in hospitals to improve air flow. We found that ventilation needs to be improved to prevent airborne transmission (Lu et al, 2021) and ventilation can be optimised by simple interventions such as keeping doors on the corridor side closed and frequently ventilating with windows (Park et al, 2023) or placing patients in beds on opposite sides of a ward first rather than next to each other, to minimise aerosol transmission (Li et al, 2007).

HEPA filtration was shown to be effective in multiple studies. Butler et al (2023) measured particulate matter (PM) levels throughout a ward before and after activation of a HEPA/UV-C air-cleaning unit and found its activation to significantly reduce PM levels. Additionally, these levels were reduced throughout the ward, not solely near the device (Butler et al, 2023). Mousavi looked specifically at optimising the location of portable air purifying units and found that it is best placed near the patient’s bed (Mousavi et al, 2020). Similarly, Conway Morris et al (2021) detected COVID in the air before activating their HEPA + UV-C unit, but not once it was in use (Conway Morris et al, 2022), Oberst also came to the same conclusion (Oberst & Heinrich, 2021). All the findings suggest portable air filtration devices can improve patient and healthcare worker safety by reducing airborne transmission. This suggestion was supported by the finding that transplant patients treated in HEPA-filtered rooms experienced lower incidences of pneumonia than those in rooms without HEPA filtration (Vokurka et al, 2014).

In addition to their clear clinical benefits, HEPA filters are easy to deploy and cost effective. Buising et al (2014) recommends further that doors to patient rooms be kept closed while these devices are in use, to optimise air cleaner function (Buising et al, 2022). Lastly, Fennelly stresses that a combination of natural ventilation and HEPA-filtration is more effective than either method alone. This study found that an air filtration unit further increased clearance of airborne particles, but that the best clearance rate came from this device being on and the windows being open (Fennelly et al, 2023). Therefore, best reduction of airborne transmission can be achieved by maximising the efficacy of natural ventilation and air-filtration devices.

Three different devices lead to improvements or reductions in health conditions, ventilator-assisted pneumonia decreased after the implementation of an eUVGI in a HVAC system (Ryan et al, 2011), invasive aspergillosis incidences decreased after installing a HEPA filter (Abdul-Salam et al, 2010) and decreases in non-invasive ventilation, nebuliser use and intubation after using a portable PECO air purifier (Rao et al, 2020).

## Need for further research

The authors that examined the efficacy of natural ventilation called for further studies investigating the effect of the addition of air purifiers (Park et al, 2023). The studies that investigated the combined effect of natural ventilation with air purifiers such as HEPA filters and UV sterilisation suggested exposure to real viral particles such as SARS-COV-2 (Oberst & Heinrich, 2021) and measuring the presence or absence of infection in healthcare professionals and patients as an outcome (Conway Morris et al, 2022). Conway Morris et al (2022) also called for an assessment of the potential harm of adding air purifiers to medical wards, through effects on noise, reduced ambient humidity, and impact on the delivery of care (Conway Morris et al, 2022). Studies that used ACUs suggest investigating their effect in larger rooms with assessments of multiple small ACUs vs. a small number of higher flow rate ACUs, they also suggest examining the best placement of these units to maximise clearance of air and particles.

Ryan et al (2011) suggested that DNA testing would definitively link HVAC and NICU environmental reservoirs with patient’s organisms, they suggested this be done in a formal RCT and would therefore, provide comprehensive findings.

## Strengths and limitations

This review was strengthened but also limited by time restrictions, search terms and the number of databases searched. These are key features of rapid reviews allowing for swift reports, although only four databases were searched they are central databases for the research conducted. This review was also strengthened by having three reviewers searching for articles and cross-checking the relevance of peer-reviewed articles. The MMAT was used to assess the quality of the included publications, this was difficult for the majority of studies which were classed as quantitative descriptive. A lot of the MMAT questions were not appropriate for the included studies, this makes it difficult to be certain of their quality. The eight evaluation studies were a strength of this review, with most evaluations having several conditions to allow comparison between the groups. Articles that also included pre-and post-intervention groups effectively highlighted the impact and difference specific units or methods made.

## Conclusions

There are numerous methods currently being used in hospitals to improve airflow, air cleaning units (ACUs), natural ventilation, low pressurised rooms, ACUs with HEPA filters or a combination of methods. Eight studies provided evaluation results and suggest that a combination of the methods listed above, usually ACUs, HEPA filters and natural ventilation are the most effective methods to reduce PM levels. Papers identified that the air change rates currently in use were not often enough to ensure effective air filtration of the room and to maintain the filters effectiveness, their efficiency must be monitored regularly. Finally, articles also outlined the importance of air flow in hospitals to reduce infections, identifying that using HVAC, HEPA or ACUs does improve patient outcomes, in terms of infection but also in nebulisation, intubation and non-invasive ventilation.

## Data Availability

All data produced in the present study are available upon reasonable request to the authors
All data produced in the present work are contained in the manuscript

## Abbreviations

ACH: Air change per hour
ACU: Air cleaning unit
AFU: Air filtration unit
eUVGI: Enhanced ultraviolet germicidal irradiation
HEPA: High efficiency particulate air
HSCT: Hematopoietic stem cell transplantation
HVAC: Heating ventilation and air conditioning
IA: Invasive aspergillosis
ISO: Isolation room
NICU: Neonatal intensive care unit
PAC: Precision air conditioning
PECO: Photo electrochemical oxidation
PM: Particulate matter
SARS-CoV-2: Severe acute respiratory syndrome coronavirus 2

## Appendices

### 1) Search Strategy

#### Themes

##### #1 Outcome terms

Exp Virus Diseases/ OR Exp Respiratory Tract Infections/ OR “respiratory infection*”.ti,ab. OR “respiratory virus*”.ti,ab. OR “respiratory tract infections”.ti,ab. OR Exp Pneumonia/ OR Exp COVID-19/ OR Exp SARS-CoV-2/ OR Exp Coronavirus/ OR Coronavirus*.ti,ab. OR “Coronavirus infection*”.ti,ab. OR 2019-nCoV.ti,ab. OR 2019 ncov.ti,ab. OR nCov.ti,ab. OR Covid19.ti,ab. OR SARSCoV-2.ti,ab. OR “novel coronavirus”.ti,ab. OR “novel corona virus”.ti,ab. OR covid*.ti,ab. OR “severe acute respiratory syndrome”.ti,ab. OR “coronavirus 2”.ti,ab. OR “coronavirus disease”.ti,ab. OR “corona virus disease”.ti,ab. OR “new coronavirus”.ti,ab. OR “new corona virus”.ti,ab. OR “new coronaviruses”.ti,ab. OR “novel coronaviruses”.ti,ab. OR Sars.ti,ab. OR “sars corona virus”.ti,ab. OR “respiratory infectious disease*”.ti,ab. OR “acute respiratory disease*”.ti,ab. OR “influenza like illness”.ti,ab. OR Exp Pandemics/ OR Pandemic*.ti,ab. OR “respiratory disease”.ti,ab.

##### #2 Intervention terms

(“filtrate”[All Fields] OR “filtrated”[All Fields] OR “filtrates”[All Fields] OR “filtrating”[All Fields] OR “filtration”[MeSH Terms] OR “filtration”[All Fields] OR “filtrations”[All Fields] OR (“recirculate”[All Fields] OR “recirculated”[All Fields] OR “recirculates”[All Fields] OR “recirculating”[All Fields] OR “recirculation”[All Fields] OR “recirculations”[All Fields]) OR (“ventilated”[All Fields] OR “ventilates”[All Fields] OR “ventilating”[All Fields] OR “ventilation”[MeSH Terms] OR “ventilation”[All Fields] OR “ventilate”[All Fields] OR “ventilations”[All Fields] OR “ventillation”[All Fields]) OR “natural ventilation”[All Fields] OR “heating”[All Fields] OR “HVAC”[All Fields] OR ((“mechanical”[All Fields] OR “mechanically”[All Fields] OR “mechanicals”[All Fields] OR “mechanics”[MeSH Terms] OR “mechanics”[All Fields] OR “mechanic”[All Fields])

AND “systems”[All Fields]) OR “system”[All Fields] OR “system s” [All Fields] OR “systems”[All Fields])) OR “Airconditioning”[All Fields] OR ((“ventilated”[All Fields] OR “ventilates”[All Fields] OR “ventilating”[All Fields] OR “ventilation”[MeSH Terms] OR “ventilation”[All Fields] OR “ventilate”[All Fields] OR “ventilations”[All Fields] OR “ventilator s”[All Fields] OR “ventilators, mechanical”[MeSH Terms] OR (“ventilators”[All Fields]

AND “mechanical”[All Fields]) OR “mechanical ventilators”[All Fields] OR “ventilator”[All Fields] OR “ventilators”[All Fields] OR “ventillation”[All Fields]) OR ((“indoor”[All Fields] OR “indoors”[All Fields]) (“airflow”[All Fields] OR “airflows”[All Fields]) OR (“aerosol s”[All Fields] OR “aerosolic”[All Fields] OR “aerosol generating procedures”[All Fields] OR “aerosol generating procedure”[All Fields] OR “aerosolization”[All Fields] OR “aerosolizations”[All Fields] OR “aerosolize”[All Fields] OR “aerosolized”[All Fields] OR “aerosolizer”[All Fields] OR “aerosolizes”[All Fields] OR “aerosolizing”[All Fields] OR “aerosols”[MeSH Terms] OR “aerosols”[All Fields] OR “aerosol”[All Fields]) OR ((“airborn”[All Fields] OR “airborne”[All Fields]) AND (“precaution”[All Fields] OR “precautions”[All Fields]))) AND (y_1[Filter])

##### #3 Environment terms

Hospitals [tiab] OR Clinic [tiab] OR Clinics [tiab] OR Infirmary [tiab] OR “healthcare facilities” [tiab] OR “healthcare facility” [tiab] OR “health care facilities” [tiab] OR “health care facility” [tiab] OR “medical centre” [tiab] OR “health centre” [tiab] OR “emergency department” [tiab] OR ED [tiab] OR “accident and emergency” [tiab] OR “a and e” [tiab] OR Exp hospital* OR “emergency room” [tiab] OR Ward [tiab] Strategy

For the initial exploration of results to determine sensitivity versus breadth we are looking at: 1 + 2 + 3

##### Final strategy and search

##### #1 Outcome terms

“respiratory infection” OR “respiratory virus” OR “respiratory tract infections” OR Pneumonia OR COVID-19 OR SARS-CoV-2 OR influenza

##### #2 Intervention terms

“HEPA filters” OR HEPA OR “UV-C systems” OR UV-C OR “laminar air flow systems” OR “filtration” OR “recirculation” OR “airflow”

1 + 2

Date range: No time restriction

##### Website searching

1. The Cochrane Library: 1867 results
2. Web of Science: 3344 results
3. MEDLINE: 1298 results
4. Inclusion Criteria

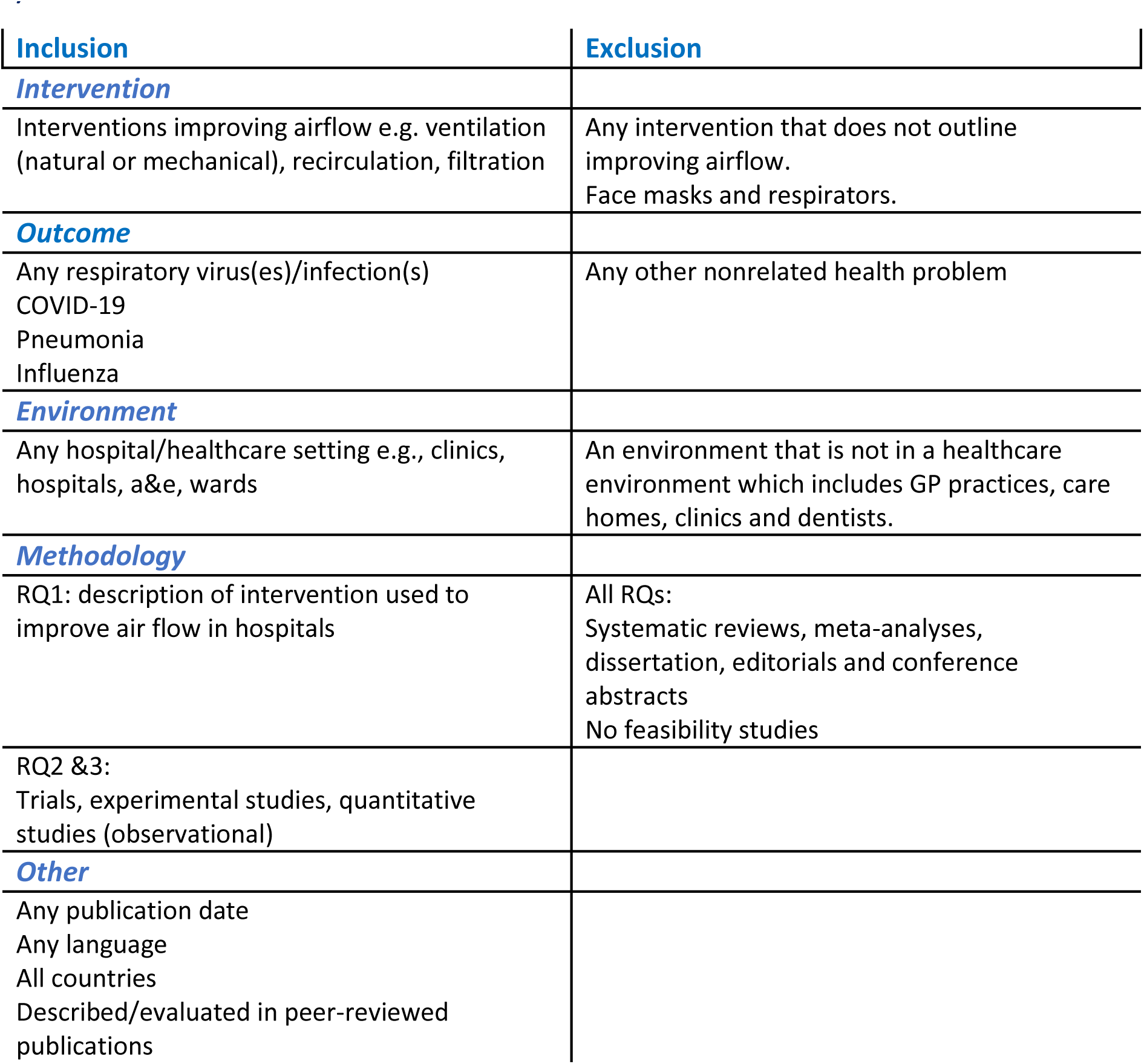

2) Inclusion Criteria
| Inclusion | Exclusion |
| --- | --- |
| **Intervention** |  |
| Interventions improving airflow e.g. ventilation (natural or mechanical), recirculation, filtration | Any intervention that does not outline improving airflow.<br>Face masks and respirators. |
| **Outcome** |  |
| Any respiratory virus(es)/infection(s)<br>COVID-19<br>Pneumonia<br>Influenza | Any other nonrelated health problem |
| **Environment** |  |
| Any hospital/healthcare setting e.g., clinics, hospitals, a&e, wards | An environment that is not in a healthcare environment which includes GP practices, care homes, clinics and dentists. |
| **Methodology** |  |
| RQ1: description of intervention used to improve air flow in hospitals | All RQs:<br>Systematic reviews, meta-analyses, dissertation, editorials and conference abstracts<br>No feasibility studies |
| RQ2 &3:<br>Trials, experimental studies, quantitative studies (observational) |  |
| **Other** |  |
| Any publication date<br>Any language<br>All countries<br>Described/evaluated in peer-reviewed publications |  |

